# Clinicians’ perspectives and experiences of providing cervical ripening at home or in-hospital in the United Kingdom

**DOI:** 10.1101/2022.12.20.22283722

**Authors:** Cassandra Yuill, Mairi Harkness, Chlorice Wallace, Helen Cheyne, Mairead Black, Neena Modi, Dharmintra Pasupathy, Julia Sanders, Sarah J Stock, Christine McCourt

## Abstract

Induction of labour, or starting labour artificially, is offered when the risks of continuing pregnancy are believed to outweigh the risks of the baby being born. In the United Kingdom, cervical ripening is recommended as the first stage of induction. Increasingly, maternity services are offering this outpatient or ‘at home’, despite limited evidence on its acceptability and how different approaches to cervical ripening work in practice. There is also a paucity of literature on clinicians’ experiences of providing induction care in general, despite their central role in developing local guidelines and delivering this care. This paper explores induction, specifically cervical ripening and the option to return home during that process, from the perspective of midwives, obstetricians and other maternity staff. As part of process evaluation involving five case studies undertaken in British maternity services, interviews and focus groups were conducted with clinicians who provide induction of labour care. The thematic findings were generated through in-depth analysis and are grouped to reflect key points within the process of cervical ripening care: ‘Implementing home cervical ripening’, ‘Putting local policy into practice’, ‘Giving information about induction’ and ‘Providing cervical ripening’. A range of practices and views regarding induction were recorded, showing how the integration of home cervical ripening is not always straightforward. Findings demonstrate that providing induction of labour care is complex and represents a significant workload. Home cervical ripening was seen as a solution to managing this workload; however, findings highlighted ways in which this expectation might not be borne out in practice. More comprehensive research is needed on workload impacts and possible lateral effects within other areas of maternity services.

## Introduction

Around one-third of pregnant women and people undergo induction of labour (IOL) in the United Kingdom (UK). Rates have risen in recent years, and they vary considerably between maternity services, with some as high as 50%, [1,2]. IOL, or starting labour artificially, is offered when the risks of continuing pregnancy are believed to outweigh the risks of the baby being born. Cervical ripening (CR) is recommended as the first stage of IOL for most women, during which a pharmacological (usually prostaglandin) or mechanical (balloon catheter or osmotic dilator) agent is applied to a woman’s cervix to cause softening, effacement and/or dilation [3]. CR stimulate labour, although further intervention such as artificial rupture of membranes (ARM) or intravenous oxytocin infusion is generally also necessary.

Increases in IOL rates come at a time of significant pressure on UK NHS maternity services, and outpatient, sometimes also referred to as ‘home’, CR has been proposed as a potential solution to manage workload and improve women’s experience [4-6]. During home CR, women undergo application or insertion of the CR agent in hospital and, following observation, return home. They then return to the maternity unit after a defined period, usually 12-24 hours, or earlier, if labour begins prior to that.

Despite the widespread implementation of home CR in the UK, there is limited evidence on safety and acceptability, impacting development of local guidelines, clinical decision-making and women’s choice. As a result, the offer of home CR is inconsistent both within and between institutions and varies in terms of the types of agents used, length of time spent at home prior to reassessment and indications and contraindications for offering this option at all [1]. There is also a paucity of literature on clinicians’ experiences of providing IOL in general, despite their central role in developing local guidelines and delivering this care, and how different approaches to CR work in practice. This paper explores IOL, specifically CR and the option to return home during that process, from the perspective of midwives, obstetricians and other maternity staff.

## Methods

### Design

This research was undertaken as part of the CHOICE Study, a prospective cohort study and process evaluation investigating cervical ripening at home or in-hospital [7]. The process evaluation (qCHOICE) involved qualitative case studies of NHS maternity units taking part in CHOICE and a postnatal questionnaire-based survey of women who had experienced IOL. Five NHS trusts and health boards in England and Scotland, selected for variation in service context and configuration, setting and method of CR, took part in qCHOICE as case study sites.

Qualitative and survey data, including relevant identifying information, were collected from clinicians, key stakeholders (e.g. Maternity Voice Partnership members), women and birth partners between November 2020 and May 2022. This paper presents an analysis of the data from clinicians, specifically midwives, obstetricians and other NHS staff. The experiences of stakeholders, women and birth partners and survey data will be reported elsewhere. Ethics approval was obtained from the National Research Ethics Service Committee (Yorkshire and the Humber—Sheffield Research Ethics Committee, REC reference: 20/YH/0145) and received National R&D approval in Scotland (NHS Research Scotland Permissions) and England (Health Research Authority).

### Data collection

Semi-structured interviews and focus groups were conducted with clinicians working in the five qCHOICE case study sites, as well as an additional NHS maternity unit participating in CHOICE. Sampling was purposive, and we anticipated undertaking between ten interviews and three focus groups comprising six to eight participants in each site. Recruitment was conducted remotely through local Principal Investigators, and we approached clinical directors, heads of midwifery, service managers, midwives, obstetricians, other NHS staff, such as pharmacists, to take part. Due to COVID-19 restrictions, all interviews and focus groups were conducted remotely online by CY, MH and CW. The interviews focused on implementation of local CR guidelines in practice, experiences of providing IOL care more generally, views on acceptability of home CR from a service perspective, and facilitators and barriers to offering at home CR safely. All interviews were video or audio recorded and then transcribed in full before being imported to a bespoke NVivo 12 database to support analysis. An abductive approach was employed for the thematic analysis [8,9]. This involved an iterative process of analysing of themes emergent in the data in relation to existing theory, knowledge and ideas in order to make further connections and insights not previously evident.

#### Descriptive findings

Table 1 provides an overview of the case study sites, including their CR pathways, as reported in local guidelines and by participants during data collection.

**Table 1.** Overview of the case study sites, including their CR pathways.

|  | Site 1 | Site 2 | Site 3 | Site 4 | Site 5 |
| --- | --- | --- | --- | --- | --- |
| **Type of unit** | Obstetric unit and midwifery unit (alongside) | Two obstetric units and two midwifery units (alongside) | Obstetric unit | Obstetric unit and two midwifery units (freestanding) | Obstetric unit and midwifery unit (alongside) |
| **Geographical location** | England (Mid) | England (South) | England (North) | Scotland (East) | Scotland (West) |
| **Area type** | Urban inner city | Suburban | Mixed urban and rural | Mixed urban and rural | Mixed urban and rural |
| **IOL rate** | 35-45% | 45% | 55-60% | 33% | 34% |
| **IOL information first given** | 35-37 weeks | 34-36 weeks | 38 and 40 weeks | 38-39 weeks | 38-39 weeks |
| **Membrane sweeps offered** | 40 weeks | 38 weeks | 1-2 per week with >48 hours in between | 40 weeks | 40 weeks |
| **Gestation IOL offered (low-** | 41-42 weeks (41+3 suggested) | 41-42 weeks (timing not specified) | 41-42 weeks (timing not | Offer at 40+7 IOL booked at 40+12 | Offer at 40+7 IOL booked at 40+12 |
| risk<sup>ii</sup>/prolonged pregnancy) |  |  | specified) |  |  |
| **Who offers IOL** | Midwives (low-risk, PROM at term) and obstetricians (high-risk) | Midwives and obstetricians (high-risk) | Midwives (low-risk) and obstetricians (high-risk) | Midwives (low-risk) and obstetricians (high-risk) | Midwives (low-risk, shared high-risk) and obstetricians (high-risk) |
| **Offer home CR** | Yes | No (suspended) | Yes | Yes | Yes |
| **Who does CR?** | Midwives | Midwives (Prostaglandin)<br>Obstetricians (Balloon catheter) | Midwives (Prostaglandin, low-risk) | Midwives | Midwives (Prostaglandin, some Balloon catheter)<br>Obstetricians (Balloon catheter) |
| **Home CR eligibility** | Low risk post-dates pregnancy | N/A | Low risk post-dates pregnancy | All unless significant concern about pregnancy | Low risk post-dates pregnancy |
| **CR methods offered** | Prostaglandin<br>Dilapan-S during SOLVE | Prostaglandin (parous)<br>Balloon catheter | Prostaglandin<br>Dilapan-S | Prostaglandin<br>Balloon catheter | Prostaglandin<br>Balloon catheter |
|  | Trial [10] | (nulliparous) |  |  | Foley catheter |
| **Methods for home CR** | Prostaglandin | N/A | Dilapan-S | Balloon catheter | Balloon catheter |
| **Protocol for those who defer or decline CR** | Individualised management plan made by Consultant Obstetrician.<br>Offer of twice weekly ultrasound, membrane sweeps, ultrasound estimation of maximum amniotic pool depth | Consultant Obstetrician appointment and management plan. Offer of increased monitoring (twice weekly CTG, growth and Doppler scan) | Offered increased antenatal monitoring consisting (at least twice-weekly CTG, ultrasound estimation of maximum amniotic pool depth) | Counselling with a “senior clinician”.<br>Offered increased antenatal monitoring (at least twice weekly CTG, ultrasound estimation of liquor volume) | Referral to senior obstetrician for risk discussion and documentation. Increased monitoring (twice weekly CTG, weekly scan for LV and umbilical artery dopplers) |
| **Where does CR take place (if not home)** | Eight-bed IOL suite or four-bed bay on antenatal ward | Unit 1: Dedicated bay on antenatal ward.<br>Unit 2: Specific room allocated for CR on antenatal ward. | Single ensuite rooms on labour suite | Antenatal ward (Maternity assessment area) | Antenatal ward dedicated bay and single rooms |
| **Women's journey during hospital CR** | Admitted to IOL suite for assessment and start of CR.<br>Transfer to labour ward when able to ARM, in labour or clinical reason. | Admitted to antenatal ward for assessment and start of CR. Transfer to labour ward when able to ARM, in labour or clinical reason.<br>Unit 2: Stay on antenatal ward for 24 hours then move up to labour ward. If they had CR by balloon and are not in labour, they have Prostaglandin for an additional 6 hours before moving to labour ward. | Admitted to IOL suite for assessment and start of CR.<br>Transfer to labour ward when able to ARM, in labour or clinical reason. | Admitted to antenatal ward for assessment and start of CR. Transfer to labour ward when able to ARM, in labour or clinical reason.<br>After 24 hours (if not in labour) are reassessed and receive Prostin or ARM. | Admitted to antenatal ward for assessment and start of CR.<br>Transfer to labour ward when able to ARM, in labour or clinical reason. If using Cook's balloon ARM must be performed within 2 hours of removal. |
| **Women's journey during home CR** | Attend IOL suite for assessment, monitoring and start of CR. Returns home for up to 24 hours. | - | Attends IOL suite for assessment, monitoring and start of CR. Returns home, with plan to return at an agreed time. | Attends antenatal ward for assessment, monitoring and start of CR. return home. Returns home for up to 24 hours. Balloon removed and either progress to ARM or return home to await ARM appointment time. If ARM not possible, Prostin and then stay in hospital. | Attends antenatal ward for assessment, monitoring and start of CR. Returns home for up to 24 hours. If using Cook's balloon, ARM must be performed within 2 hours of removal. |

All five case study sites (Table 1) initially provided home CR; however, their local guidelines changed due to COVID-19 restriction, with one site increasing this option and another one site suspending it completely for a majority of the study period. The local guidelines for induction including methods used for cervical ripening, both at home and in hospital, and eligibility criteria for those offered the option to go home varied between sites. We conducted 45 interviews and four focus groups with midwives, obstetricians, other NHS staff and stakeholders between November 2020 and December 2021 (Table 2).

**Table 2.** Numbers of clinicians who took part in the study, by professional group.

| Method | Participant group | Number |
| --- | --- | --- |
| Interviews (n=45) | Midwives | 29 |
|  | Obstetricians | 14 |
|  | Other NHS staff | 2 |
| Focus groups (n=4) | Midwives | 8 |
|  | Obstetricians | 20 |
| Total |  | 73 |

### COVID-19 impact

During the data collection period maternity services were affected by the impact of COVID-19. This included staffing pressures, concerns about infection in hospital settings, visiting restrictions and, in some services, limitations on partners accompanying women during antenatal, labour and postnatal care.

#### Thematic findings

The thematic findings were generated through in-depth analysis of clinicians’ interviews and focus groups and are grouped to reflect key points within the process of providing IOL, specifically CR, care: ‘Implementing home cervical ripening’, ‘Putting local policy into practice’, ‘Giving information about induction’ and ‘Providing cervical ripening’.

### Implementing home cervical ripening

Most clinicians who took part were positive about home CR, perceiving it to be better for themselves in terms of managing maternity workload and also for women and their families, especially those who were deemed low-risk and healthy. Motivation for introducing home CR was stated as reducing unnecessary time in hospital for women, linked to the perception that women would prefer to be at home and that their experience would be improved by offering home CR:

> Our understanding and perception is that it will be a lot easier to manage the stress and the anxiety associated with that delay if somebody is at home and in their own environment and relaxed rather than stuck in a hospital. (Site 2 MW INT 109)
>
> [T]hey’re in their own house, aren’t they? When they come in, you go in to see them and they’re on the bed and you think, “You’re not actually sick”. They’re pinned to the beds, and they need to be up on their feet. That gravity brings that baby’s head down. If they go home even if they’re just pottering around, walking to the toilet, making a cup of tea” (Site 3 OB INT 017)

Clinicians involved with developing local IOL guidelines that include home CR discussed the significant groundwork, knowledge-building and confidence needed. Clinicians drew on their colleagues’ experience and information from other NHS trusts and boards to establish their own local guideline. For Site 4, this groundwork had taken years to build and implementation was moving slowly, until an outside factor, COVID-19, precipitated a rapid change:

> “[W]hen we got to March and COVID arrived and we didn’t want people in the hospital and women didn’t want to be in the hospital, that was the environment for change was there.” (Site 4 MW INT 075)

However, pandemic-triggered change towards home CR was not common across the case study sites and, in another site, by contrast, home CR was suspended. Many staff wanted to offer home CR but described changing guidelines as lengthy and problematic, including ensuring women were properly informed about the new service:

> “[W]omen hadn’t been given that information prior to them being admitted to antenatal ward then they’d already had it in their heads that they were coming in, so they weren’t that keen for that outpatient induction. So, you see how one if one element of the chain wasn’t performed, it had an impact on the whole how to implement the service.” (Site 5 MW INT 096)

### Putting local policy into practice

Participants were asked to talk about their local IOL policy or guidelines during interviews and focus groups, which revealed intricate care pathways involving many different actors and places (Fig 1).

**Figure 1:**
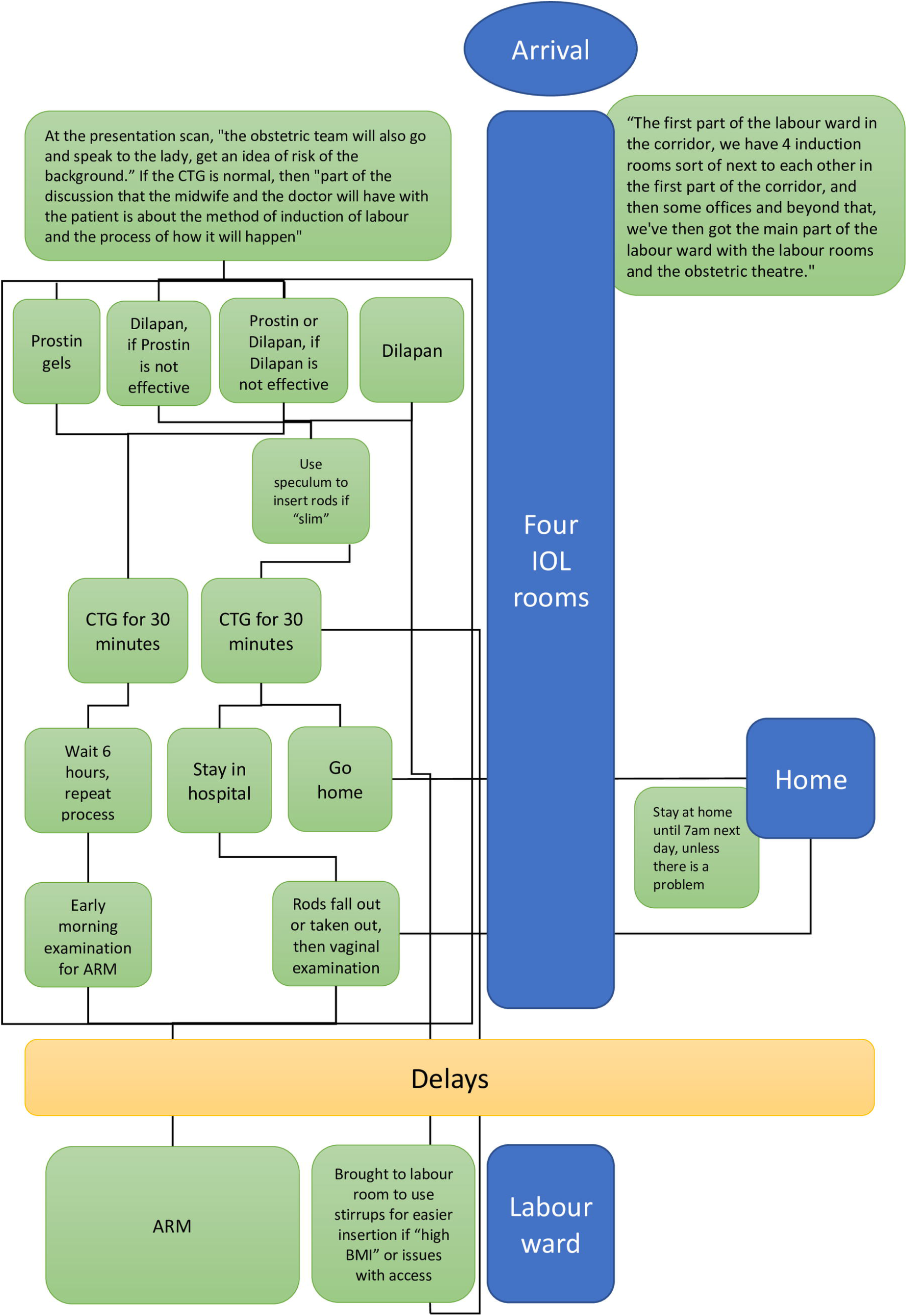
The CR process of one of the case study sites, as described during an interview with an obstetrician.

Though every case study site had a detailed guideline for providing IOL, during the interviews, we identified deviations from these. Deviations were particularly noted when new guidelines were implemented:

> “[N]ot everybody interprets that guideline the same way… The policy’s changed and not everybody’s on the same page. Things filter down and not everybody is privy to the information.” (Site 3 MW INT 044)

A midwife at another site highlighted the lack of local protocol clarity in practice stemming from issues with communication:

> “I just don’t think it’s clear. I think the biggest problem with our trust is not clear -- you’re always like its hearsay whether you find something out and you’re learning from someone else that’s telling you something and you don’t necessarily know if what they’re telling you is correct. You just kind of hope it is.” (Site 1 MW INT 022)

Policy dissemination beyond hospital into the community was described as uneven, ever changing, difficult to access and lost in a sea of communication. Others touched on the complexity of implementing a new CR guideline into practice:

> “[W]e just culturally have used inpatient induction for so long… I think the process is new and…I think new things scare people a bit.” (Site 5 OB INT 110)

In most sites, midwives were expected to take on new roles with home CR, including the responsibility of inserting catheters or osmotic dilators. Some participants identified training related to these roles as a sticking point, while midwives did not always perceive this to be the case:

> [W]e have been kind of chipping away at it for a while and trying to get enough interest, and I think that the main issue was getting enough midwives comfortable enough to put these devices in… (Site 4 OB INT 077)
>
> I was going to be providing the training for the midwives because we wanted to be midwifeled. It isn’t until you see how it’s done yourself and what are the challenges faced by a midwife that you can truly understand what the midwife and what the woman has to go through for the induction procedure… There were some midwives who were very, very keen to get it up and running. When we were all set, the guideline was all set and it was going to be an outpatient induction procedure, the pandemic had an impact on staffing… (Site 5 MW INT 096)

Some participants spoke of the different professional perspectives on IOL, including senior or junior midwives and obstetricians. For example, a non-clinical manager stated:

> There’s a very much a split in terms of views about induction between the obstetricians and the midwives. I think the obstetricians very much see induction of labour as a positive thing. …The consultants see that as a core part of their work, and they’re the ones who are very focused on improving the pathway, making sure delays don’t happen. I think there is more resistance from the midwives -- I wouldn’t say they’re resistant. That’s not the right word. I think they see induction in a different way. (Site 3 HCP INT 013)

Data from participants directly involved in providing IOL care instead reflected tensions arising from different roles in the process, including gaps between an ideal of how home CR might work and the everyday reality in their service.

### Giving information about induction

Ensuring women are able to make informed decisions about their options is stated to be essential during maternity care, yet participants often spoke of a mismatch in perspectives between themselves and women undergoing IOL. Managing expectations using realistic information about the process and expected duration of IOL was viewed as crucial for closing the gap between these perspectives:

> “[W]omen become frustrated with how long it takes, how long it goes on for, so I’ve decided that the best way to manage it is to give them a timeline of what can happen. It’s just managing their expectations of it because in the past we’ve had bad feedback from women not really knowing what was happening. The other thing that I’ve found is their expectations of care during the induction process. You know, they’re not ill. They don’t need anyone, there’s no sudden danger, and they ought to just be left, so what I try to explain is treat this room like a little hotel room. This is your room. This is an ordinary day.” (Site 3 MW INT 019)

This midwife, for example, was critical of unrealistic messaging around the process, revealing the paradoxical nature of women being told they are at risk, in need of an immediate IOL, and then providing care in which they are treated as ‘low-risk’ for a prolonged period:

> If you get someone sensible who books their induction, they’ll have told them this can take up to a week. If you get a doctor who can’t be arsed and just wants to see them in and out to clinical triage, they go, “Oh yeah, there’s a risk of stillbirth. We need to get your baby out. I really think you need to be induced as soon as possible. We’ll book you in for tomorrow. Goodbye.” That’s the end of the conversation. The women come, they think they’re going to have the baby in their arms by the next day. (Site 1 MW FG 027)

While clinicians recognised the importance of being proactive in providing information on all the dimensions of IOL, many reported this process as uneven and varied from person to person. This was described as primarily due to time constraints:

> “The community midwives don’t have a lot of time. They have a 20-minute appointment, and that’s to do the woman’s normal antenatal check and then discuss if she wants to have one (IOL) and then discuss induction of labour with them.” (Site 3 MW INT 018)

Resources constraints, in the form of tools and evidence, were also identified by participants as obstacles to effective information giving and risk communication:

> “I think we lack the tools to do that because we’re having this discussion. We’re still only doing it verbally. There are no good infographics. There’s definitely nothing in a non-English language to help women to understand this really quite complex thought process and assimilation of evidence but also lack of evidence and appreciating uncertainty and weighting of evidence.” (Site 1 OB INT 050)

### Providing cervical ripening

The decision-making process around IOL is complex because the IOL pathway and its associated risks are themselves complex. IOL was typically offered and booked by midwives for those deemed low-risk and with prolonged pregnancies between 41-42 weeks’ gestation. Obstetricians offered and booked IOL for those deemed high-risk, and the timing of IOL for these groups varied between sites (Table 1), with several local guidelines recommending provision be individualised. Generally, the logistics of the CR process were managed by midwives, but their role in applying the CR agent varied between sites, often based on which one was being used (Table 1).

Delays were an issue throughout the IOL process and affected women having home and hospital CR. Delays frequently occurred between the CR procedure and admittance to labour ward for ARM, some reportedly lasting upwards of seven days. This incurred further delays in admitting women for CR and administering a second CR agent, if needed. Participants most frequently attributed delays to inadequate staffing:

> “There’s some where you’ve got a bed available and we walk the woman round to the room and get her in the room but we haven’t actually done anything. And then the emergency buzzer goes and clearly, we can’t carry on with the induction. So, we walk them back round to induction suite and we say, sorry, we can’t do you.” (Site 1 MW INT 025)

Some midwives described a ‘waiting list’ and calling women during the night when beds became available:

> If the woman on top of the list, we ring her, and she doesn’t answer the phone at 1 o’clock in the morning, and you think “OK, what if she rings me back at 3 o’clock and I’ve brought the next one in and she’s delayed again?”. But it’s a process -- it’s a system, and we have to follow it. If I’ve got capacity right now, she needs to come in. I will ring her three times if I have to, and I’ll document that, and I’ll move on to the next one. If the next woman doesn’t pick up, then I’ll move on to the next one. I’ll keep going until I’ve got them all in. (Site 1 MW FG 031)

IOL was recognised by many as having a large impact on midwives’ workload, requiring additional planning and communication with women. Besides the day-to-day logistics of having more women than staff could care for, midwives also described being on working groups to improve issues caused by IOL. One participant (non-clinical) regarded IOL as planned work and a potential solution to workload issues; however, maternity care is unpredictable, and this can limit the capacity to provide IOL:

> There will be delays unfortunately. I think just to manage it as best we can, but then the unpredictability of our workload hugely impacts on the ability to offer induction. (Site 4 MW INT 067)

Clinicians discussed their staffing shortages and how this contributed the delays and their ability to provide IOL care. While some suggested home CR would create a its own set of logistical issues, most believed it would be a solution to the delays in and dissatisfaction with the IOL process:

> “We do have at the staffing issues, I’m sure quite a lot of hospitals do. I mean, I’m down to one ward at the moment, normally have two so if these women can stay at home safely, then it’s much better for us than have them just sitting here looking at us thinking, is it me next?” (Site 4 MW INT 090)

Despite this perception, the rates of home CR varied between the case study sites, and many were not offering home CR widely in practice. Sites where it was only available to those who are ‘low-risk’ with prolonged pregnancies had few women going home. This, coupled with restriction on methods used, limited the number of women being offered and having home CR. Site 4, where home CR was regarded as a safe option for most women and established in their local guidelines, was the only exception. Clinicians reported home CR as part of routine IOL care, with around “80%” of women going home.

Establishing safety, especially in relation to using different CR methods, within a context of limited evidence was cited as a concern. Though the NICE guidance at the time of the study did not recommend mechanical methods for IOL [11], they are regarded as the safest by staff:

> I am a bit nervous about a home induction with Propess, just because of the way we can see some women react to it. I would be comfortable with home induction if we used Dilapan or Foleys. I think most midwives would be comfortable with that. Home induction with Propess makes me a little bit twitched as to what, if anything, goes wrong and they haven’t realised, and they’re not being monitored, are they? (Site 1 MW INT 020)

Most sites only used mechanical CR methods for home, though some saw downsides to these. They were reported as difficult to insert and painful for women, requiring an ARM within a relatively short period after removal, often regarded as unachievable due to unit capacity. As with much of the rest of the IOL process, clinicians had a range of views on CR methods, as to which was more effective or least painful:

> I remember one or a couple of nights every single woman on the induction bay had been given Dilapan, and it was great. They all slept through the night, no one was having any contractions or in pain. A couple of little niggles here and there which a bit of paracetamol would sort out. And they all woke up ARMable. (Site 1 MW INT 022)
>
> [W]e have been using Cook’s balloon, but I think women were finding it more uncomfortable, so you have to provide more analgesia…I think it was just generally more uncomfortable than they [women] were with the Prostin. They [clinicians] seem to be getting better results with them [Cook’s balloons]… (Site 5 HCP INT 064)
>
> I don’t know if it’s the position of where the balloon was when it was inserted, but I’m finding more and more patients are needing Prostin. I mean, from your conversations with others, have you found that, that’s there’s failure to have the ARM when the balloon comes out? (Site 2 MW INT 032)

There were also conflicting views on the impact of home CR on workload; some felt it decreased workload, while others did not. Women still come to hospitals for CR insertion and monitoring, they call the hospital for advice, and they return after 12-24 hours, or if they go into labour or have complications. Midwives described additional workload from managing women in different places and the stress of not having staff and beds available for them to return:

> [I]t is acknowledged as a risk because with the staffing levels as they are at the moment, we can only get a woman to a certain point in that induction process, if we haven’t got a midwife available to do that and give them the one-to-one care on Labour Ward, to be able to break the waters and get on with it. Once the amniotomy is done, obviously, we have to look after them, and if we don’t have staff, then we can’t, and we end up with delay. Over the last couple of months, it’s not uncommon to see delays of 24 hours plus with women just sitting in a ward waiting for that to happen, which, as I said earlier, I’m aware of the fact that with our staffing levels it may not completely remove that risk by doing an outpatient induction. (Site 2 MW INT 109)

## Discussion

Our research has demonstrated that providing IOL care is complex, and that home CR is being implemented with varying success across the NHS, given the uncertainties that surround it, including gaps in the evidence base and unpredictable nature of intrapartum maternity care work. We recorded a range of practices and views related to IOL, showing how the integration of home CR is not always straightforward and how deviations from local guidelines might occur when put into practice. Variations in practice are consistent with an earlier survey we conducted on IOL practice changes due to COVID-19, which revealed differences in responses even from clinicians working in the same NHS trust [1]. Moreover, the differences between and within local guidelines could also be a contributing factor to the variation in IOL rates across trusts and boards [2].

Most clinicians interviewed were positive about home CR and spoke of the benefits of women being at home during CR. They perceived it as a preferable option for women and their families, despite limited evidence to date on women’s experiences of home CR [12,13]. Superficially, home CR appears to be about improving women’s experiences, but it is also strongly linked to managing workload. It was largely seen as a solution to the many “operational challenges” of IOL care; however, this perspective is based on under-developed and, at present, inconclusive evidence about the safety and efficacy of home CR or the impact on workloads and resources. Revising an IOL guideline and implementing home CR involves many actors from across the maternity service, and hinges, in part, on the extent to which a maternity service makes this the default option as well as the confidence and engagement of clinicians working both in community and hospital. Staffing issues and changes in clinicians’ roles were key obstacles identified; however, the latter was not reported as a barrier ubiquitously across all professional groups taking part in this study.

Midwives were often responsible for CR, and with home CR, they are expected to take on new responsibilities of inserting catheters or osmotic dilators, requiring training and additional workload. Some midwives saw downsides to those methods – they are difficult to insert, painful for women, and require an ARM within a relatively short period after removal, which was often unachievable due to delays. Research has shown catheters can be painful and require skill to insert [12] but further studies are needed regarding timing and ARM use. Others perceived mechanical agents to be safer for home CR, and their use has now been included in the updated NICE guidance [13]. The ‘low-risk’ inductions were expected to be midwife-led, including providing information, doing the CR insertion, managing the processes of sending women home or keeping them in hospital, responding to queries or calls, handling readmissions, triaging and doing ARMs.

Clinicians who took part in this study were frequently looking for strategies to manage their induction workload, and home CR was one such proposed solution. Yet, they also highlighted ways in which this expectation might not be borne out in practice. Home CR did not necessarily lessen workload but rather shifted it from one space, or person, to another and, in some cases, increased it. Some participants perceived this as due to “unpredictability” of maternity; however, IOL itself is unpredictable. CR, in particular, is acknowledged to be a “lengthy” process, requiring “numerous resources” [6], including repeated monitoring and assessment and often further intervention in the form of more CR agents, ARM and intravenous oxytocin infusion. Unlike planned caesarean sections, IOL can take days, compounded by delays due to staff shortages. Delays were acknowledged as anxiety-inducing for women, and during these periods there was often a mismatch between clinicians’ and women’s understanding of levels of risk and expectations for ‘pre-labour’ care. As part of qCHOICE, we interviewed both women and their birth partners, and their experiences will be reported in a subsequent publication.

Our findings highlight services’ limited preparation for increased IOL rates, in terms of workload changes and additional staff time. Initiatives aimed at reducing stillbirth, such as the implementation of the Saving Babies’ Lives Care Bundle (SBLCB), did indicate a potential increase in IOL rate, yet there has been little elaboration to date on the wider effects this would have on services. The SPiRE study, which evaluated the implementation of SBLCB, took a narrow view of service impacts, focussing on more on economic analysis and clinical outcomes than on lateral effects, such as delays [14]. However, it found 19.4% increase in IOL at sites that implemented SBLCB and identified that “additional resources” needed for “secondary demands” connected to bundle, including IOL, were a potential barrier [14 pp xvi]. In addition, the AFFIRM trial found a significant increase in IOL rates associated with a care package for fetal movement monitoring, although it did not find a significant reduction in stillbirth [15]. With the increasing numbers of IOL that will likely be midwifery-led and involve home CR, more comprehensive research is needed on the workload impacts of these approaches and possible ripple effects within other areas of maternity services.

Throughout the IOL process, from giving information to providing CR, time was limited and delays were frequent. Clinicians reported efforts to keep the process moving, even if this meant calling women repeatedly in the middle of the night when staff and space became available. This ‘process-production’ approach to care, recalling anthropological and sociological work on the use of industrial models and mechanistic metaphors in maternity [16-18], responds to pressure on staff and services but disregards the wellbeing of women, physically and psychologically. Women need to fit with the “system”, or they miss out, passed over for the next person on the list. This approach sits in opposition to national maternity policy ideal of person-centred and compassionate care; however, it was a way for staff to cope with an unmanageable workload connected to increasing IOL rates.

We argue that this process-production approach, coupled with the expectations of home CR, are indicative of a trend towards routinisation of IOL in maternity care. As IOL increases and its related indications expand, a paradoxical concept of the ‘low-risk induction’ has emerged. Women are deemed ‘low-risk’ enough to “just be left” or sent home during induction, which has been advised because they or their babies are perceived to be at risk. Despite the pervasiveness of risk in maternity care [19], once the first stage of induction begins, women and babies are in “no sudden danger”, and risk is treated as all but eliminated. There are, however, risks associated with IOL, such as hyperstimulation [3], because it remains an intervention into the physiological course of pregnancy. For clinicians in the study, induction was part of an “ordinary day”, though research suggests that for women it is anything but ordinary [12]. As we have shown, IOL is not without lateral effects. At the current rates, IOL already represents a significant workload in UK maternity services, which, at the time of this study and of writing, are experiencing staffing shortages so acute that induction care is reported as incurring additional risks. In light of this, making IOL more routine, whether at home or in-hospital, should be reconsidered, and its potential risks, including unintended effects and those beyond the usually recognised clinical scope, require further scrutiny.

## Data Availability

The data presented in this research article has not been made open access due to ethical concerns about confidentiality and identification of participants. There are limited numbers of clinicians working in induction of labour care in the United Kingdom, meaning indirect identification is more likely, even if the data set is de-identified. Given the nature of qualitative research, complete anonymisation of the data, in which all direct and indirect identifying information is removed, is not possible. Moreover, the data set contains potentially identifying and sensitive patient information. For these reasons, open access has been restricted to protect the confidentiality of participants and identities of patients.
The data set is available on request. Requests may be made to National Research Ethics Service Committee, specifically the Yorkshire and the Humber Sheffield Research Ethics Committee at, using REC reference 20YH0145.

## Acknowledgements

We are grateful to those who gave their time for interviews and focus groups despite the severe workload pressures and ongoing COVID-19 pandemic.

CHOICE is funded by the National Institute of Healthcare Research Health Technology and Assessment (NIHR HTA) NIHR 127569. SJS is funded by a Wellcome Trust Clinical Career Development Fellowship (209560/Z/17/Z). The funders had no role in study design, data collection and analysis, decision to publish, or preparation of the manuscript. The views expressed are those of the authors and not necessarily those of the National Institute of Healthcare Research or the Department of Health and Social Care.

## Notes

We use ‘pregnant women and people’ when speaking more generally about maternity and care in recognition of the diverse gender identities of those who become pregnant, experience IOL and give birth. However, the respondents in this study used ‘woman’ and ‘women’ exclusively, so our reporting reflects this usage.

“Low-risk” and “high-risk” are used in this table to reflect the wording frequently used in site guidelines.

